# Effect of deep brain stimulation in patients with severe and treatment-resistant obsessive-compulsive disorder

**DOI:** 10.1101/2025.11.06.25338313

**Authors:** Andrés Gempeler, Patricia Gómez Salgado, Janet Puñal Riobóo, María del Carmen Maceira Rozas, Yolanda Triñanes Pego, The Living Evidence to Inform Health Decision program (LE-IHD) research group

## Abstract

**Objective:** To provide a timely, rigorous and continuously updated summary of the available evidence on the effectiveness of deep brain stimulation (DBS) in people with severe and refractory obsessive-compulsive disorder (OCD).

**Methods:** We conducted a systematic literature review of intervention, as the baseline synthesis report of a living evidence approach. We followed the methodological framework by Elliot et al. 2014. The L.OVE platform from Epistemonikos was used for identification, screening and pre-selection of studies. Systematic reviews (SR) and randomized clinical trials (RCT) evaluating the use of DBS in patients with severe and refractory OCD were included. Data extraction and risk of bias assessment were performed in duplicate using appropriate instruments. Primary outcomes were symptom severity (efficacy) and adverse events (safety), secondary outcomes were quality of life, functionality, effect on cognitive function, patient perception and values. Evidence certainty was assessed with the GRADE approach.

**Results:** The searches identified 45 references for full text review, finally, 16 SRs and 12 crossover RCT (112 patients) were included. The pooled estimates suggested a favorable effect of DBS on reducing symptoms measured by Y-BOCS score in the short term (MD=7.3 points, 95%CI: 3.9–10.6, *p* <0.0001) and the long term (MD=14.8 (95% CI: 12.1 – 17.5) (*p* <0 . 0001), with high and moderate certainty evidence, respectively. The estimated pooled incidence of permanent serious adverse events (SAE) was 6% (95% CI: 0%–19%), and surgical SAE was 9% (95% CI: 2%–18%); both with very low certainty due to serious imprecision. Non-pooled results from individual studies also suggested benefits of the interventions on quality of life and cognitive function.

**Conclusions:** Deep brain stimulation of proposed targets for refractory obsessive-compulsive disorder has a large beneficial effect on reducing symptoms severity in the short and long term. Implantation has a low incidence of adverse events, and a high incidence of non-serious adverse events during the stimulation calibration period. Benefit to harm balance should consider uncertainty on the risk of adverse events related to the surgical procedure and to the device, and appears favorable given its large effects on symptom reduction.

## BACKGROUND

Obsessive-compulsive disorder (OCD) is a neuropsychiatric condition characterized by unwanted obsessions and/or compulsions (1) . With a prevalence of 1-3% in the general population, OCD represents a common mental disorder that has a significant impact on quality of life, generating disability and a relevant burden for patients and their families (2, 3) .

First-line treatment for OCD involves the use of selective serotonin reuptake inhibitors (SSRIs) and/or cognitive behavioral therapy (CBT) with exposure and response prevention (4–8) . However, it has been reported that between 10% and 40% of patients may not respond to these conventional treatments and may suffer severe and persistent symptoms (4) .

For this group of patients with severe and refractory OCD, ablative neurosurgery would be indicated. In recent years, the role of other less invasive interventions, such as deep brain stimulation (DBS) or transcranial magnetic stimulation, has been investigated. These interventions could be an alternative to consider before ablative surgery (4, 9) .

The objective of this review is to synthesize the most relevant evidence available to inform decision-making on the use of deep brain stimulation in patients with severe and refractory OCD in the Servizo Galician Health Service (SERGAS). Furthermore, a living evidence framework has been selected, as it is considered an emerging technology for which new evidence is likely to be published that can inform decision-making.

This synthesis of living evidence has been produced as part of a broader health technology assessment report under the Living Evidence Programme. to Inform Health Decisions (10), which supports healthcare organizations in implementing a living process for developing evidence syntheses to inform healthcare decisions. The ongoing update resulting from the evidence monitoring will be available on the Program website (https://livingevidenceihd.com/lesrepo/)

### Protocol and registration

This report conforms to the PRISMA guidelines ’ Preferred Reporting Items for Systematic reviews and Meta-Analyses ’ (11, 12) . The protocol of this systematic review has been published in OSF (https://osf.io/rbt98).

### Deviations from the protocol

In the review protocol, a score ≥ 28 on the Yale-Brown Obsessive Compulsive Scale was defined as a criterion of severity following some previous studies (13) . Given the variability in the identified studies, studies with patients with severe and refractory OCD were finally considered for inclusion at the discretion of the authors, following the DSM diagnostic criteria, a less restrictive definition of “treatment resistant” or “refractory” based on scores on the Y-BOCS (> 24).

## METHODS

### Design

We started a process of living evidence synthesis with this baseline synthesis according to the Living methodological framework Evidence Synthesis (LES), and the Methods for Planning and Reporting Living Evidence Synthesis proposed by Bendersky et al. (14).

### Search Strategies

The literature search was devised by the team maintaining the L·OVE platform using the following approach:

- Identification of terms relevant to the population and intervention components of the search strategy, using Word2vec technology to the corpus of documents available in Epistemonikos Database.
- Discussion of terms with content and methods experts to identify relevant, irrelevant, and missing terms.
- Creation of a sensitive boolean strategy encompassing all the relevant terms
- Iterative analysis of articles missed by the boolean strategy, and refinement of the strategy accordingly.
- Application of validated filters to identify clinical trials and non-randomized studies in the MEDLINE database.

For the baseline synthesis, the searches were covered from the inception date to April 2023 when the monitoring of new evidence started. No date, language, study design, publication status, or language restriction was applied to the searches. Table 1 presents the searches defined by the type of sources.

**Table 1.** Boolean Searches by source.

|  |  |
| --- | --- |
| **Source** | Epistemonikos Database |
| **Boolean Strategy** | ((obsess* OR compulsi* OR OCD*) AND (((("deep brain" OR "deep-brain") AND (stimulat* OR neurostimulation*)) OR DBS) AND ("critical review" OR "electronic search" OR "evidence-based analysis" OR "evidence-based review" OR "literature search" OR "meta analysis" OR "meta synthesis" OR "meta-analyse" OR "meta-analytic review" OR "meta-study" OR "meta-synthesis" OR "metaanalysis" OR "metasynthesis" OR "meta-analysis" OR "pooled effect" OR "random-effects model" OR "systematic quantitative review" OR "systematically searched" OR "systemic review" OR (review AND randomized) OR (systematic AND review) OR MEDLINE OR "literature review" OR PubMed)) |
| **Type of studies** | Systematic reviews |
| **Date of coverage** | From inception to 27/04/2023 |
| **Source** | Epistemonikos Database, Pubmed |
| **Boolean Strategy** | ((obsess* OR compulsi* OR OCD*) AND (((("deep brain" OR "deep-brain") AND (stimulat* OR neurostimulation*)) OR DBS) AND (randomi* OR RCT OR placebo* OR trial OR "controlled-trial" OR randomly*)) |
| **Type of studies** | Randomized controlled trials |
| **Date of coverage** | From inception to 27/04/2023 |
| **Source** | Epistemonikos Database, Pubmed |
| **Boolean Strategy** | ((obsess* OR compulsi* OR OCD*) AND (((("deep brain" OR "deep-brain") AND (stimulat* OR neurostimulation*)) OR DBS) AND (((("follow up" OR "follow-up" OR followup*) OR observation* OR epidemiologic*) AND (stud* OR evaluation*)) OR retrospective* OR prospective* OR longitudinal* OR observational* OR "non-randomized" OR "non-randomised" OR "non randomized" OR "non randomised" OR nonrandomi* ) OR ("regression-discontinuity" OR (regression AND discontinuity) OR kink) OR (mendelian AND randomi*) OR (multivaria* OR multinomial* OR logistic* OR regression OR cox OR stepwise* OR adjust* OR independent* OR (receiver* AND operat*) OR AUC OR ROC) OR (focus OR "focus-group" OR qualitative* OR interview* OR "grounded theory" OR "mixed-methods" OR "mixed methods" OR semistructured* OR "semi-structured" OR informant OR ("open-ended" AND question*) OR nvivo OR verbatim) OR ((test-negative OR "test negative") AND (study OR design OR controls OR "case-control")) OR (nested*) OR ((quasiexperiment* OR "quasi experimental" OR "quasi-experimental") OR "pretest-posttest" OR pretest* OR posttest* OR "pre-post" OR "pre-post tests" OR "post test" OR "post tests" OR pretest OR pretests OR "pre test" OR "pre tests" OR (repeated AND measure*) OR "time series" OR "time-series" OR "matched controls" OR beforeafter OR "before-after" OR "before after" ) OR ("interrupted-time-series" OR (interrupted AND time AND series*) OR ((ITS* OR "time-series" OR "time series") AND (study OR studies OR analys* OR design*)) OR (trend AND (time OR analys*)) OR ((segmented* OR piecewise* OR "broken-stick" OR "broken stick") AND regression*)) OR (extension AND (rct OR randomi* OR trial OR posttrial* OR "post-trial")))) |
| **Type of studies** | Primary studies with a design different from a randomized study. |
| **Date of coverage** | From inception to 27/04/2023 |

### Clinical question

Table 2 presents the clinical question in PICOD format (Population, Intervention, Comparison, Outcomes, and Design).

**Table 2.**
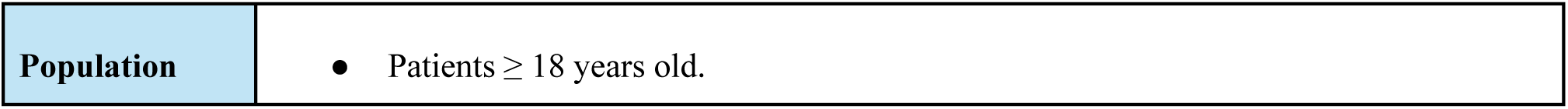
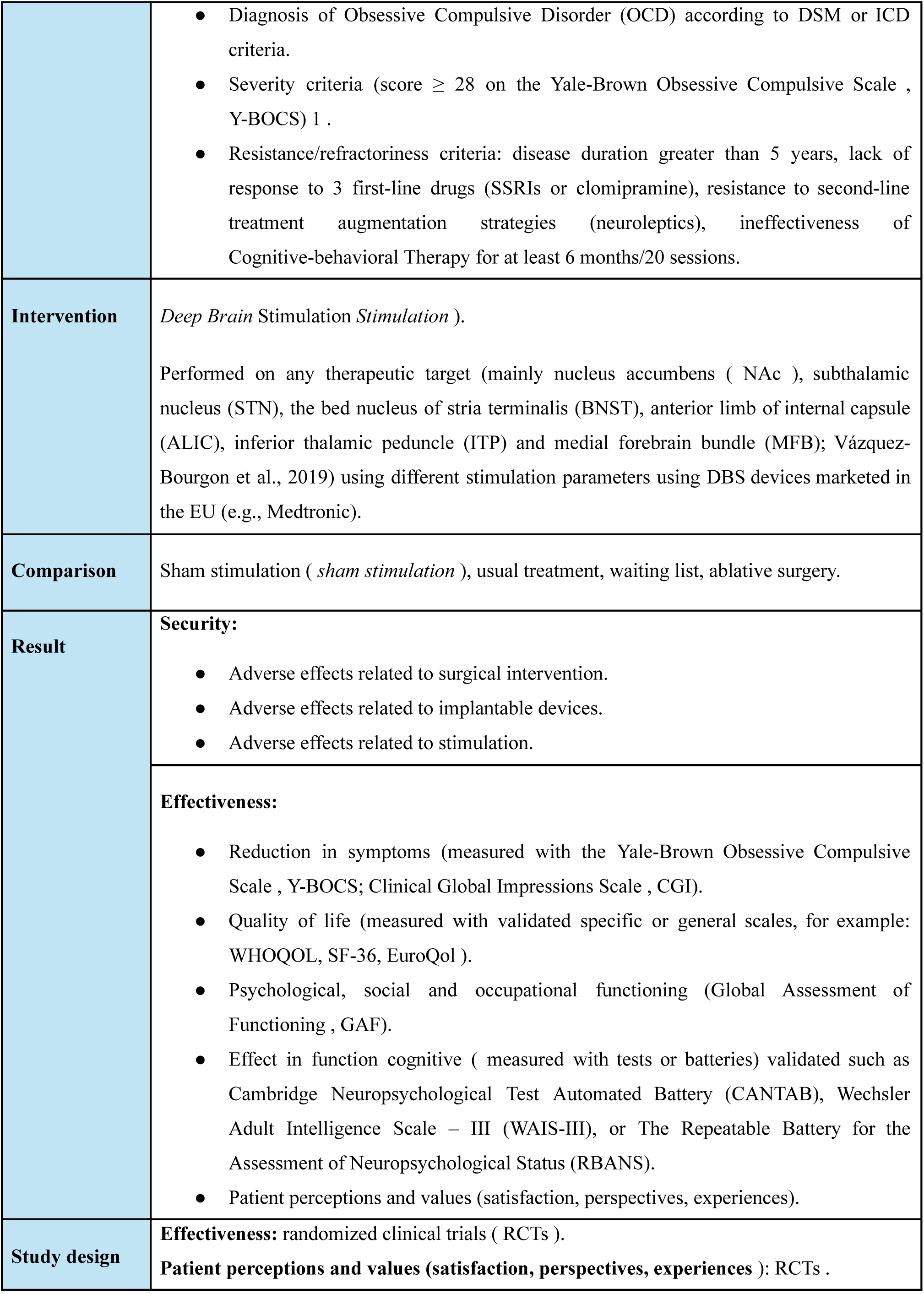
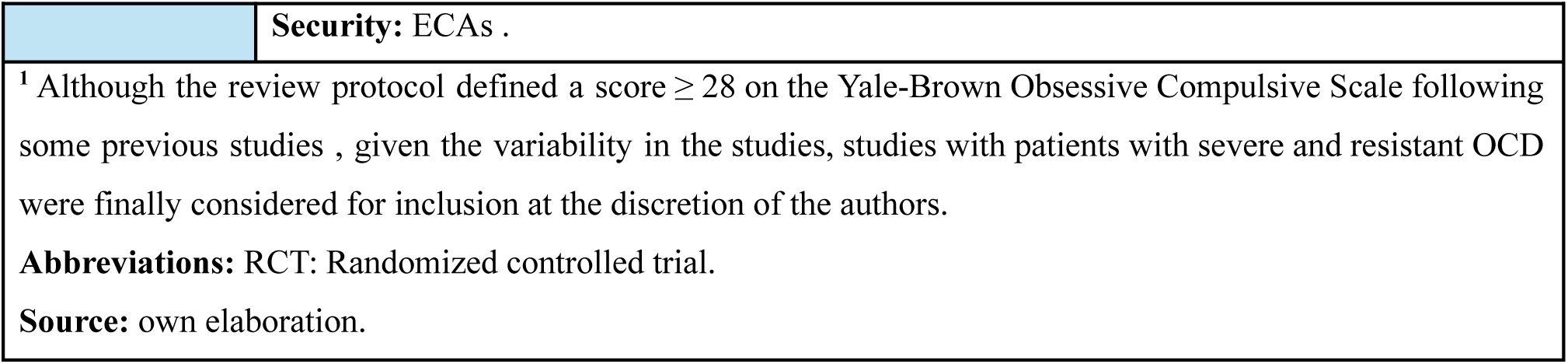
: **Clinical question in PICOD format**

Evidence identification, screening and selection

Following a preliminary search of health technology assessment (HTA) reports and clinical practice guidelines (CPGs), a specific search strategy was designed to answer the PICOD question.

The processes of identification, selection, and review of evidence were carried out with the support of technological facilitators developed by the Epistemonikos Foundation and included in its *Living Overview platform. of Evidence* (L·OVE) (40) (15) .

A continuous automated search was performed using the multiple information sources and databases maintained by the Epistemonikos database, following its procedures (41). The results of the literature searches were automatically incorporated into the L·OVE platform, specifically implemented for the question “Deep brain stimulation for treatment-resistant obsessive-compulsive disorder,” where automated classifiers excluded references with a low probability of being relevant to this question. No restrictions on date, language, study design, or publication status were applied to the searches. The *Collaboratron platform* facilitated duplicate screening and document management based on the search results.

An evidence matrix was created that compiled those systematic reviews that met the inclusion criteria along with the primary studies from which they were based, so that these reviews served as a source for identifying RCTs .

Subsequently, the search for the systematic review considered the parent of the evidence matrix was updated (from January 2021, the date the parent review strategy was implemented) with the aim of identifying recently published RCTs not included in the systematic reviews. To identify ongoing trials, the ICTRP and clinicaltrials.gov databases were searched.

### Information extraction

A reviewer extracted data from each included study, which were recorded in evidence tables. Information was collected on study design, participant characteristics (including disease severity and age), study eligibility criteria, details of the intervention and comparator, outcomes assessed and their timing, study funding source, and conflicts of interest reported by the investigators.

data for meta -analysis were either extracted directly from the primary studies (in three studies – Welter 2021, Denys 2010, Huff 2010), or calculated by the authors from individual patient data reported in the primary studies (in three other studies – Barcia 2019, Luyten 2016, Abelson 2005-), or were obtained as a result of a combination of both strategies (in three other studies – Tyagi 2019, Mallet 2008, Nuttin 2003). For one of the studies, this information was obtained through one of the published SRs that had included it (Goodman 2010).

### Risk of bias assessment

(16) was used to assess the risk of bias of systematic reviews ADDIN EN.CITE . The assessment of the risk of bias of the included RCTs was performed using the RoB 2 tool (Risk of Bias 2) (17) for the evaluation of randomized clinical trials (RCTs) with crossover trials). This assessment was performed in duplicate, and discrepancies were resolved by consensus or by a third reviewer. Tables and figures for the risk of bias of the studies were created using the Visualization web tool. RoB Tool.

### Summary of results

We performed a quantitative synthesis of the evidence using meta-analysis for each outcome. If we found differences in study characteristics that would have made their pooled analysis incorrect, we summarized the findings for each outcome narratively.

#### Meta-analysis of efficacy – Y-BOCS scale

The reduction in OCD symptoms according to the Y-BOCS scale was assessed in two ways that respond to the two-phase design (cross-over and maintenance) of the studies that have evaluated this intervention:

1. Comparison of Y-BOCS scores between ON and OFF periods during crossover phases ; considered experimental evidence of the short-term effect of the intervention.
2. Comparisons of Y-BOCS scores before (baseline) and after (stimulation), which allowed to evaluate the effect of the intervention and implantation surgery in the short and long term (BL vs On), considered observational (quasi-experimental) evidence.

The summary measures used to assess the intervention effect were the mean difference (MD) and the standardized mean difference (SMD, also known as Hedges ’ g) . We used both because the MD allows for a more useful clinical interpretation in terms of Y-BOCS score reduction. However, the SMD provides a cumulative estimate that facilitates the application of recommendations for the assessment of the certainty of the evidence (described below) .

The overall and subgroup effects were estimated using a random-effects model. Studies lacking direct data for any of the indicated time points were not included in the meta-analysis for the corresponding comparison. Subgroups based on the brain stimulation target were used to assess differences in intervention delivery and account for potential heterogeneity in outcomes across studies.

#### Meta-analysis of adverse events

For the meta-analysis of the intervention’s effect on safety outcomes, a proportional meta-analysis was used over the entire follow-up period to estimate the risk of serious adverse events (SAEs), including surgical SAEs and neuropsychiatric SAEs, which were meta-analyzed together and separately. Additionally, information reported by the studies on the resolution of SAEs was used to estimate the risk of permanent serious adverse events. The effect on these safety outcomes was assessed by subgroups according to follow-up duration (12 months, 21 months, and 4 years or more). The overall and subgroup effects were estimated using a random-effects model.

The effect of the intervention compared with non-intervention controls could not be assessed due to the nature of the studies and the heterogeneity in the definition, collection, and reporting of safety outcomes during the crossover phase of the studies.

#### Heterogeneity

Heterogeneity between studies was examined using the I^2^ statistic . calculated from the MD and SMD meta-analyses to identify heterogeneity due to differences in baseline Y-BOCS scores. Heterogeneity was classified as low (I^2^ : 0–24.9%), moderate (I^2^ : 25%–49.9%), high (I^2^: 50% –74.9 %), and very high (I^2^ :75%–100%). Calculations were performed with Review Manager 5.4.1 software and RStudio using the “ metafor ” and “ meta ” analysis packages .

For the remaining efficacy and safety variables, heterogeneity in definition, time horizon or reporting made it inappropriate to conduct meta-analysis, so a qualitative synthesis of the evidence (narrative) was chosen.

Evaluating the certainty of the evidence

The certainty of the evidence for each outcome was assessed using the GRADE methodology (Recommendations Assessment, Development and Evaluation working group methodology, GRADE Working Group) (18, 19), in the domains of risk of bias, inconsistency, indirect evidence, imprecision, publication bias, confounding, effect size and dose-effect gradients. Certainty was classified as high, moderate, low or very low.

#### Type of evidence

For the outcomes obtained from the crossover phase (On vs Off) of the study, the evidence was considered experimental, and was considered to be of high certainty at the start of the assessment. For this outcome, the risk of bias assessment derived from the RoB2 Crossover tool was used. trials .

#### Risk of bias

ON) comparison outcomes, the available evidence was considered observational due to the absence of a control arm. The consequences of the risk of bias on the certainty of the evidence for these outcomes were defined according to the validity of the measurements and patient follow-up, without being affected by some domains with risk of bias in the RoB2 Crossover tool. Trials related to randomization, washout and carry-over periods .

#### Inconsistency

To assess inconsistency between studies, we used the meta-analytic statistical measures of heterogeneity, I2, and tau. However, given the overlap of the 95% CIs with the point estimate and the lack of 95% CIs from individual studies that considered either no effect or a deleterious effect, we did not downgrade the certainty of the evidence for imprecision in the presence of high I2 or significant tau.

#### Imprecision

To assess imprecision, an optimal information size (OIS) was defined in terms of a clinically significant reduction on the Y-BOCS score. In the literature, refractory OCD is typically defined as scores above 28 points, with a maximum of 40 points. For these high Y-BOCS scores, the usual measure of reduction >35% is inadequate, as it corresponds to reductions of ∼10 points and ∼14 points for baseline scores of 28 and 40, respectively. Therefore, for severe and refractory OCD, it has been suggested that a reduction of 6 points or more be considered a clinically relevant change. For the Y-BOCS score, the standardized mean difference proposed by GRADE as the DCRM in the absence of an established clinical parameter is =0.2, which in the case of scores between 28 and 40 points results in reductions of 1 point and 0.6 points, respectively, which are not considered clinically relevant. Since a DCRM does exist in refractory OCD, we used a 5-point reduction as the minimum clinically relevant difference for the OIS calculation (point below the established 6-point threshold). This adheres to the GRADE suggestion of using a DCRM consistent with the clinical problem at hand. From large studies in patients with refractory OCD, we identified mean Y-BOCS scores of 30.8 points with a standard deviation (SD) of +/-6 points (418 patients) and 30.1 (+/-5.1) points (68 patients). With a baseline mean of 30 points and SD=5, a difference of 5 points corresponds to SMD=1.0, so the optimal information size with power=90% is 22 patients per group (44 total); for SMD=0.8 (4 points) it is 33 patients per group (66 total), and for SMD=0.6 (3 points) it is 59 patients per group (118 total). In the case of crossover or before-and-after designs, the total number of patients acts as its own control, so the information size was calculated by multiplying the total sample for each outcome by 2.

#### Publication bias

We assessed publication bias using funnel plots and Egger ’s test to evaluate the effect of small studies on efficacy outcomes.

Living Evidence Framework evidence synthesis

In order to maintain the live evidence process for this review, the Epistemonikos -L.OVE platform (15) was used as a technological enabler for evidence identification, screening, and selection. We will maintain a live search on the L-OVE platform to detect randomized controlled trials. An artificial intelligence algorithm deployed on L-OVE platform will provide instant notification of articles with a high probability of being eligible. In addition, every three months, we will manually search for ongoing studies on the WHO International Clinical Trials Registry Platform and clinicaltrials.gov.

A reviewer will be responsible for assessing the evidence submitted to L.OVE on a monthly basis and applying the selection criteria presented above. If a potentially eligible study is identified, a second reviewer will confirm its eligibility by reading the full text. The results of the evidence monitoring will be collected and stored as part of the study records. Information on PRISMA will be updated accordingly. The study selection criteria will be reviewed and amended accordingly during the LE process every four months.

All newly eligible studies will undergo data extraction. The data synthesis will be updated immediately afterward, and the body of scientific evidence for the outcomes of interest will be assessed using the GRADE approach, looking for changes in the results of the certainty assessment.

## RESULTS

### Search results

#### Systematic reviews

The initial search for systematic reviews identified 75 references. After reading the titles and abstracts, 46 were selected for full-text reading. Of these, 16 met the selection criteria (see Figure 1) and were included in the evidence matrix (see Figure 2).

**Figure 1.**
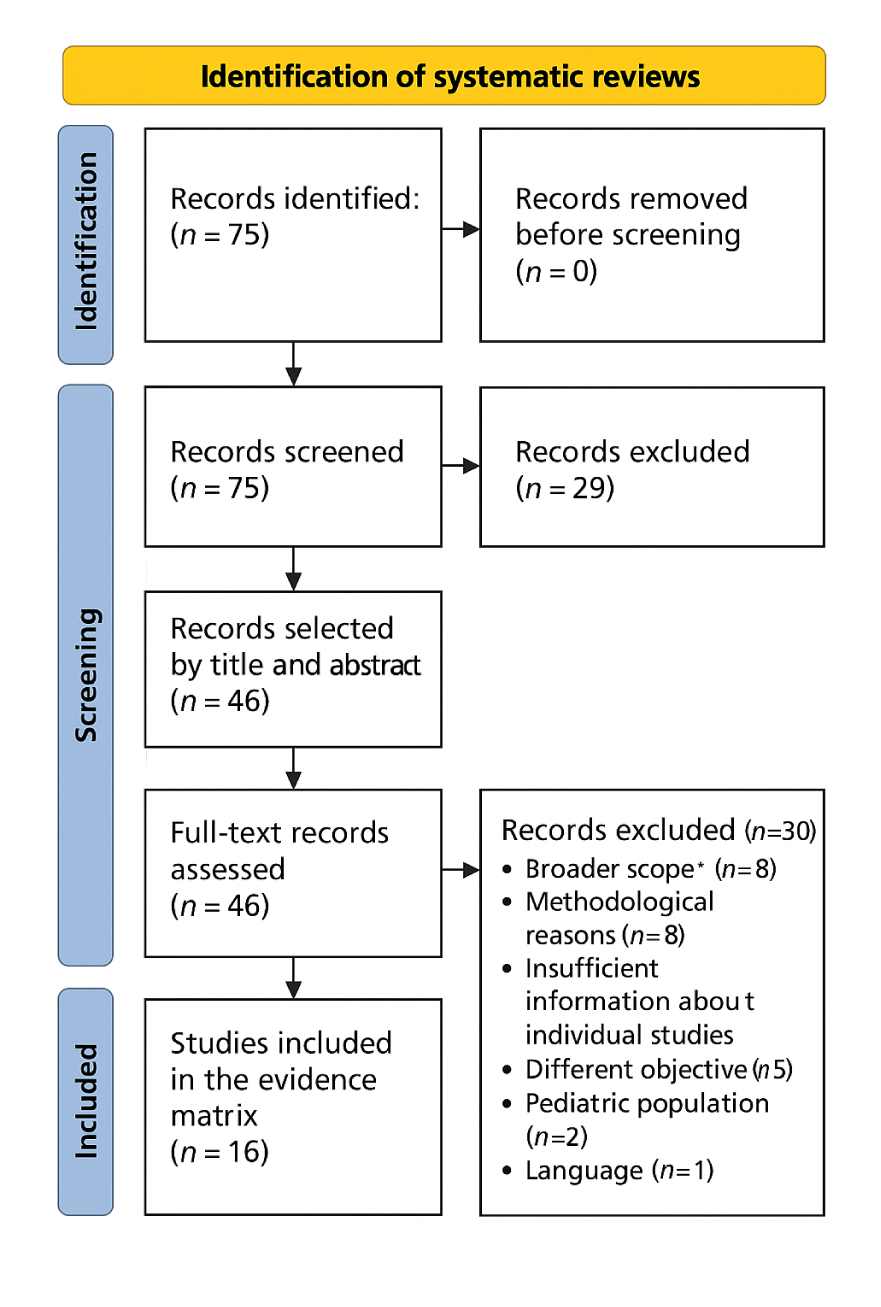
Flowchart for selecting systematic reviews. Gadot et al. (2022) (2) was selected as the parent review ; it was the most recent, the one that best fit the research question and the largest number of RCTs analyzed (n=8). Additionally, those by Mar-Barrutia et al. (3) and Martinho et al (20) were selected as reference reviews because they were also recent, and because of the number of RCTs included (n=7 and n=10, respectively, see Figure 2).

**Figure 2.**
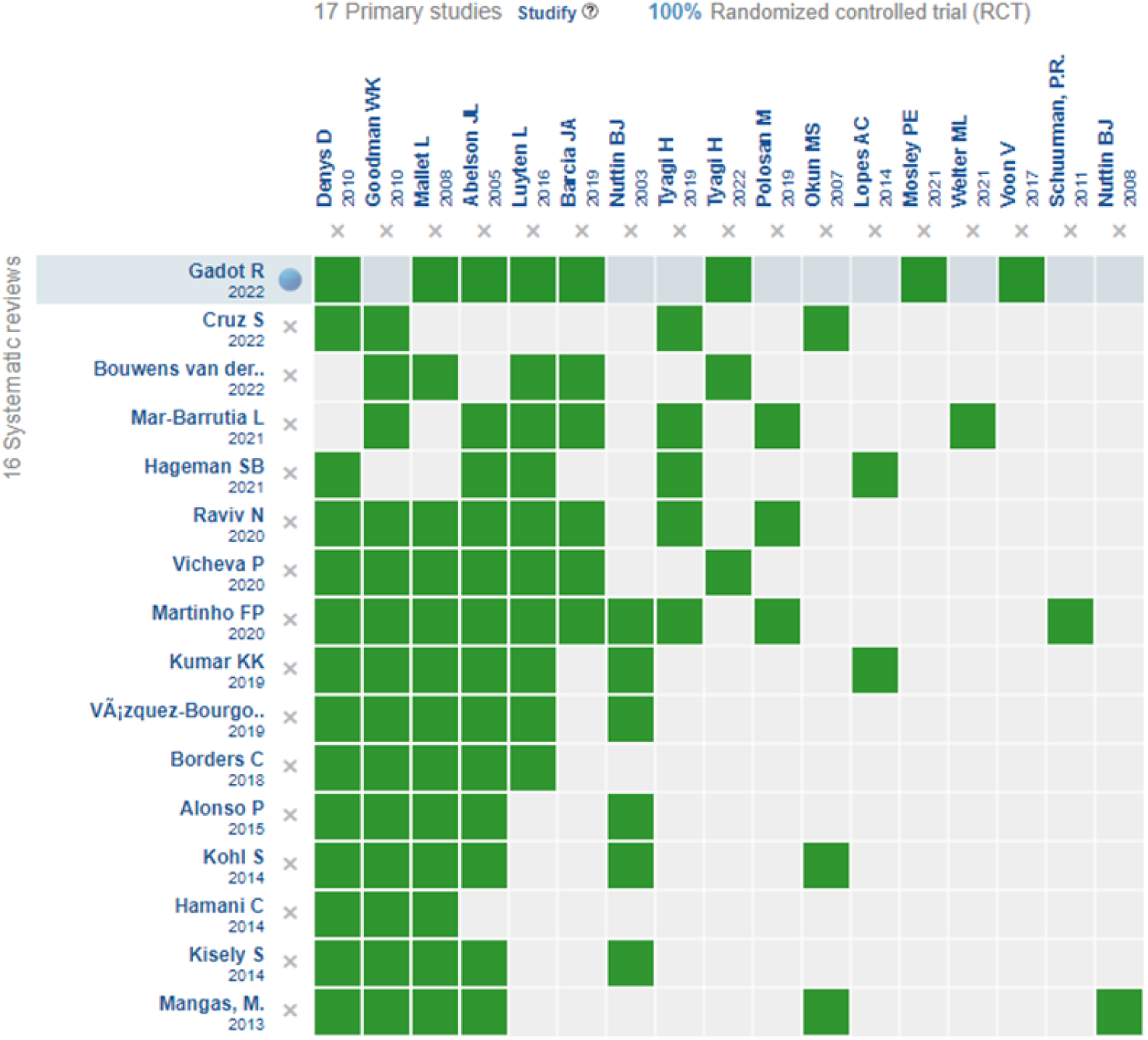
**Evidence matrix generated on the Epistemonikos platform *Source*** *: generated in the Epistemonikos platform* .

The methodological quality of the three selected reference reviews was critically low according to the AMSTAR-2 tool, mainly due to reasons such as the non-publication of the review protocol, the bibliographic search or the non-publication of the list of excluded studies.

#### Randomized clinical trials

The review matrix identified 17 eligible RCTs through 2021. Following the search strategy employed and starting from the last year searched (2021), the review matrix was updated, which identified 12 additional references. One more RCT was identified through cross-referencing, resulting in a total of 30 eligible RCT references. Twenty-three were preselected by title and abstract. Of these, 12 met the selection criteria and were included in our synthesis (see Figure 3).

**Figure 3:**
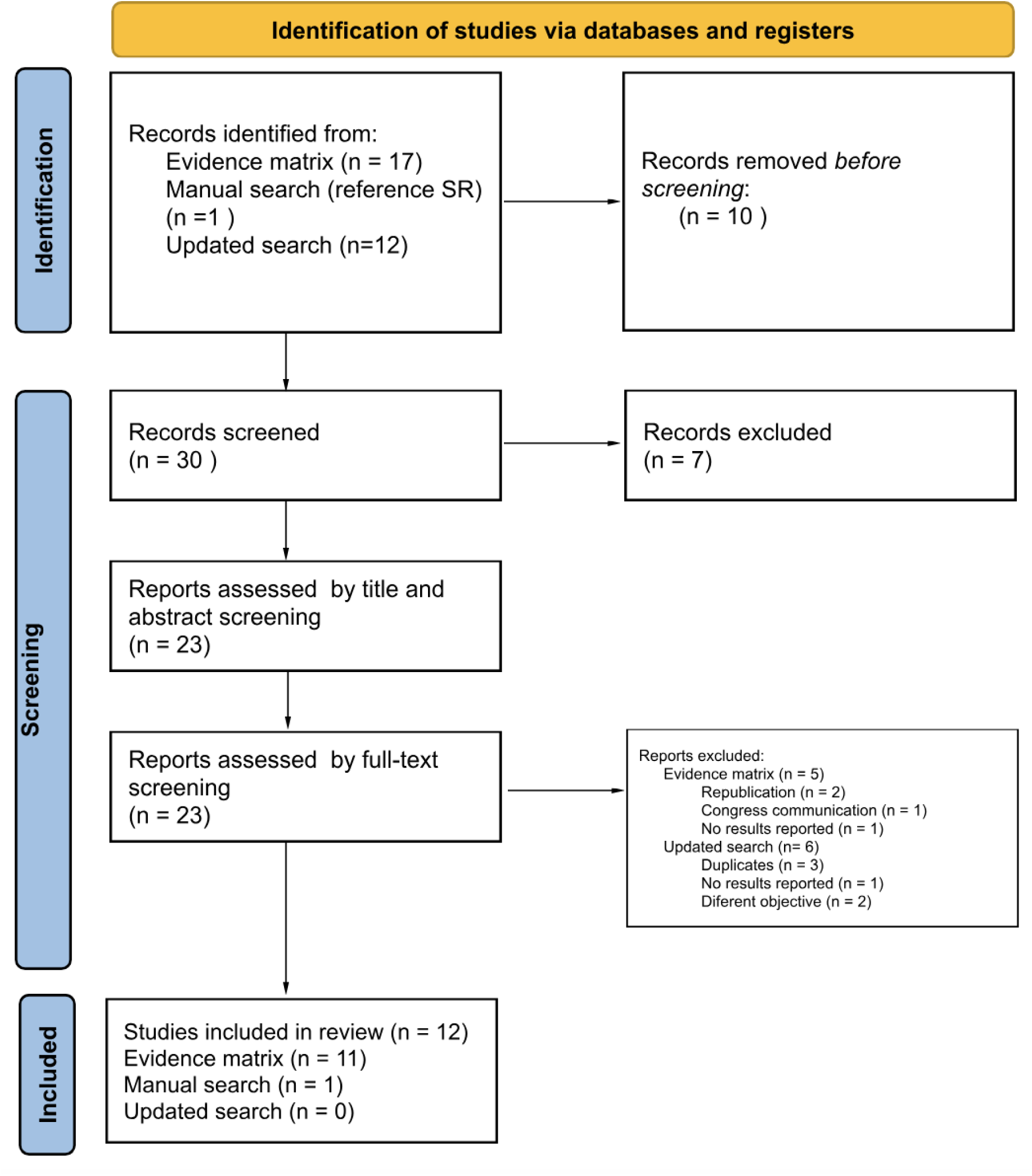
Flowchart of primary and updating studies. Most studies were conducted in Europe (3 in France (21–23), 1 in Spain (24), 1 in Germany (25), 1 in the UK (26), 1 in the Netherlands (27), 1 in Belgium (28), 1 in Belgium and Sweden (29), 2 in the USA (13, 30) and 1 in Australia (31)) . 58.3% of the studies had a sample size of less than 10 patients (range: 4 to 24 patients). Table 3 shows the characteristics of the included RCTs.

**Figure 4.**
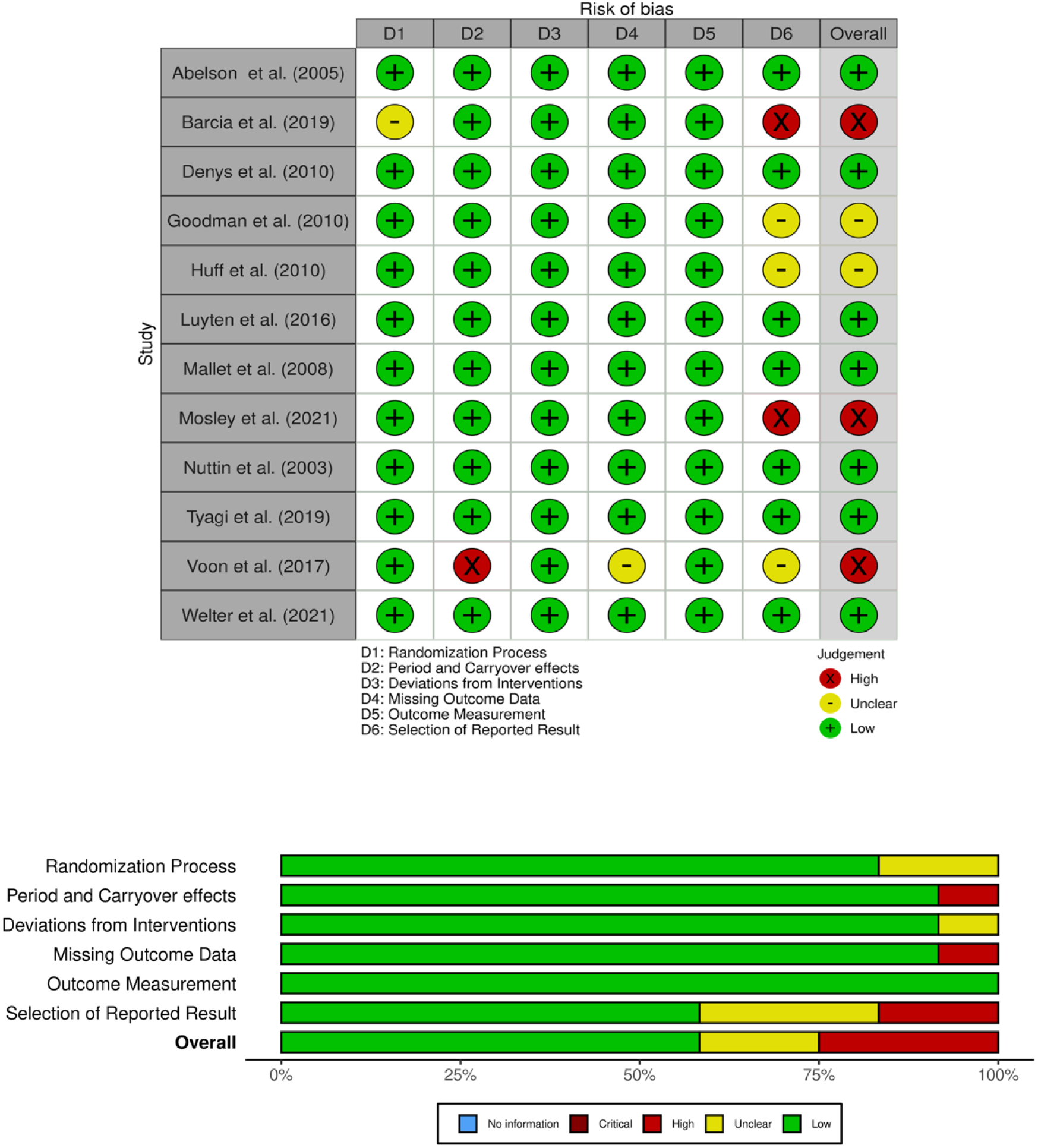
Traffic ligh charts and R oB 2.0 risk of bias summary of included studies Source. : RoB Visualization Tool

**Table 3.** Characteristics of the included clinical trials.

| Authors and year | Country | Design | No. of patients in the sample | Baseline OCD severity according to the Y-BOCS scale | Brain stimulation goal | Follow-up | Primary outcome | Complementary outcomes |
| --- | --- | --- | --- | --- | --- | --- | --- | --- |
| Abelson et al. 2005 | USA | Double-blind crossover RCT | 4 | 32.75 ± 5.85<br>Range: 26-39. | ALIC | 12.9 months<br>range: 4-23 | Change in Y-BOCS Scale score | Cognitive function |
| Barcia et al. 2019 | Spain | Crossover RCT | 7 | 32 | Nac/Sweet Spot | 2 days | - | Decisional impulsivity |
| Denys et al. 2010 | Netherlands | Double-blind crossover RCT | 16 | 33 | NAc | 21 months | Change in Y-BOCS Scale score | - |
| Goodman et al. 2010 | USA | Double-blind, stepwise RCT | 6 | 33.7 (2.42) | V CVS | 11 months | Change in Y-BOCS Scale score | Global Clinical Impressions and SF-36 |
| Huff et al. 2010 | Germany | Double-blind crossover RCT | 10 | 32.2±4.0 | NAc | 6 months | Change in Y-BOCS Scale score | - |
| Luyten et al. 2016 | Belgium | Double-blind crossover RCT | 24 | 35<br>Range 30-40 | ALIC/BST | 72 months<br>range: 15-171 | Change in Y-BOCS Scale score | - |
| Mallet et al. 2008 | France | Double-blind crossover RCT | 16 | 32.06 ±3.04 | amSTN | 10 months | Change in Y-BOCS Scale score | - |
| **Mosley et al. 2021** | *Australia* | Double-blind crossover RCT | 9 | - | ? | 12 months | Change in Y-BOCS Scale score | MADRS Scale |
| **Nuttin et al. 2003** | *Belgium and Sweden* | Double-blind crossover RCT | 4 | 34.75±3.95<br>Range 30-38. | ALIC | 18.8 months range: 3-31 | Change in Y-BOCS Scale score | Score on the Clinical Global Severity and Improvement Scales |
| **Tyagi et al. 2019** | *United Kingdom* | Double-blind compensated RCT | 6 | 36.17±0.75<br>Range 34-38 | amSTN, VCVS, Sweet Spot | 21 months | Change in Y-BOCS Scale score | Cognitive function |
| **Voon et al. 2017** | *France* | Double-blind crossover RCT | 12 | 34.3 (3.2)<br>Range 30-40 | ? | 15 months | Change in Y-BOCS Scale score | MADRS Scale |
| **Welter et al. 2021** | *France* | Double-blind crossover RCT | 8 | 33.5±4.31 | Nac, amSTN | 19 months | Change in Y-BOCS Scale score | - |

The 12 RCTs included 122 patients. Fifty percent of the studies included 50% or more women (50–87%). 33% of the studies included approximately 44% women. Nuttin et al. (29) did not report information on the sex of the included patients. The mean age of the included patients was between 36 and 47.9 years. Two studies reported the age range of the included patients (Tyagi et al. (26) : 37–62 years and Mallet et al. (21) : 29–56 years). Nuttin et al. (29) did not report information on the age of the included patients. Baseline OCD severity assessed by the Y-BOCS scale was reported in 91% of the studies with a mean score ranging from 32 to 36 points. Baseline OCD severity was not reported in 1 study (13, 21, 31) (Mosley et al.).

Table 3 summarizes the main characteristics of the included studies. All studies had a crossover design, in which patients in the intervention group remained on active stimulation (ON) for a period and those in the control group on sham stimulation (OFF) for a predetermined period. Subsequently, patients on active stimulation switched to sham stimulation, and vice versa. Some studies (21, 24, 27, 31) reported a 1-month washout period between the two ON/OFF treatment periods. In 8 of the reviewed studies (21, 23, 26–31), patients received DBS for periods ranging from 1 week to 8 months prior to inclusion in the clinical trial phase with the aim of exploring the type of stimulation, in terms of the therapeutic target, frequency, voltage or amplitude to be applied subsequently.

Eighty-three percent of the studies reported the type of device used, all of which used different models from the Medtronic brand. The characteristics of the DBS received by patients, both among different studies and within the same study, varied greatly in terms of stimulation frequency, pulse width, and voltage. Most studies (10/12) reported that the follow-up period for patients on DBS ranged from 3 to 15 months. Voon et al (22) evaluated patients for 4 hours after the intervention, while Mosley et al (31) indicated the time intervals at which they evaluated the stimulation, but not the total follow-up period.

#### Studies in progress

Ongoing studies were reviewed, of which 25 met the inclusion criteria. Of these 25 studies, 11 are in unknown status, 8 are active and not recruiting patients, and 6 are currently recruiting patients. The sample size of the studies that are recruiting patients is 2-64 patients and the countries in which they are being conducted are Israel, China (n=2), France and the USA (n=2).

### Risk of bias of the included studies

#### Effects of the intervention

##### Short-term Y-BOCS score reduction (Off vs On)

For the evaluation of the effect of sham stimulation vs. active stimulation (OFF vs. ON) on OCD symptoms according to the Y-BOCS scale, it was possible to meta-analyze the results of 7 studies (13, 21, 25, 27–30) with a total of 71 patients. Three studies (23, 24, 26) did not report information about periods of OFF sham stimulation, and a further study (31) reported a statistically significant OFF vs. ON score difference in favor of stimulation (ON) but did not report extractable data for meta-analysis . These four studies were not included in the meta-analysis of this outcome.

The pooled estimated mean difference in Y-BOCS score was a 7.3 point reduction (95 %CI: 3.9–10.6, *p* <0.0001) in favor of active stimulation (ON) with moderate heterogeneity (I2 ^=^ 37 %). In the standardized mean difference (SMD) meta-analysis (see Figure S1) the effect of stimulation on the reduction in Y-BOCS score was SMD=0.83 (95%CI: 0.47; 1.18), with low heterogeneity (I2=10%). The decrease in the heterogeneity measure with the use of SMD reflects that the heterogeneity of the studies is largely explained by differences in the baseline severity score of the patients between studies, which when standardized decreases inconsistency in the effects of the intervention . Figure 5 shows the summary of this meta-analysis .

**Figure 5.**
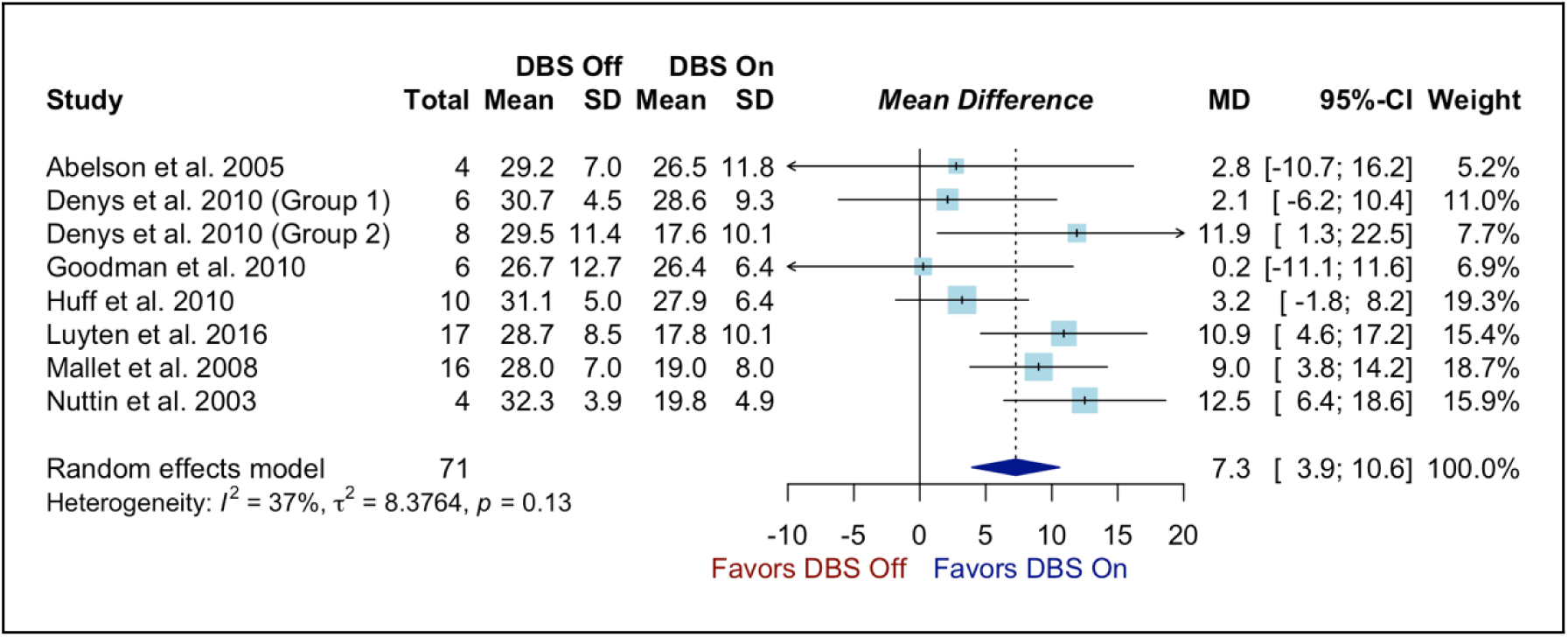
Forest plot for meta-analysis of Off vs On brain stimulation. Egger ’s regression to assess the small study effect did not detect publication bias (t = -0.23, p = 0.82) (see Figure S2). The certainty of the evidence on this effect is high, because this is experimental evidence with a low risk of bias (studies with a high risk of bias did not report information on this outcome and are therefore not included in this meta-analysis), and we did not identify any other findings that decrease the certainty of the estimate (see Table 4. SoF).

**Figure 6.**
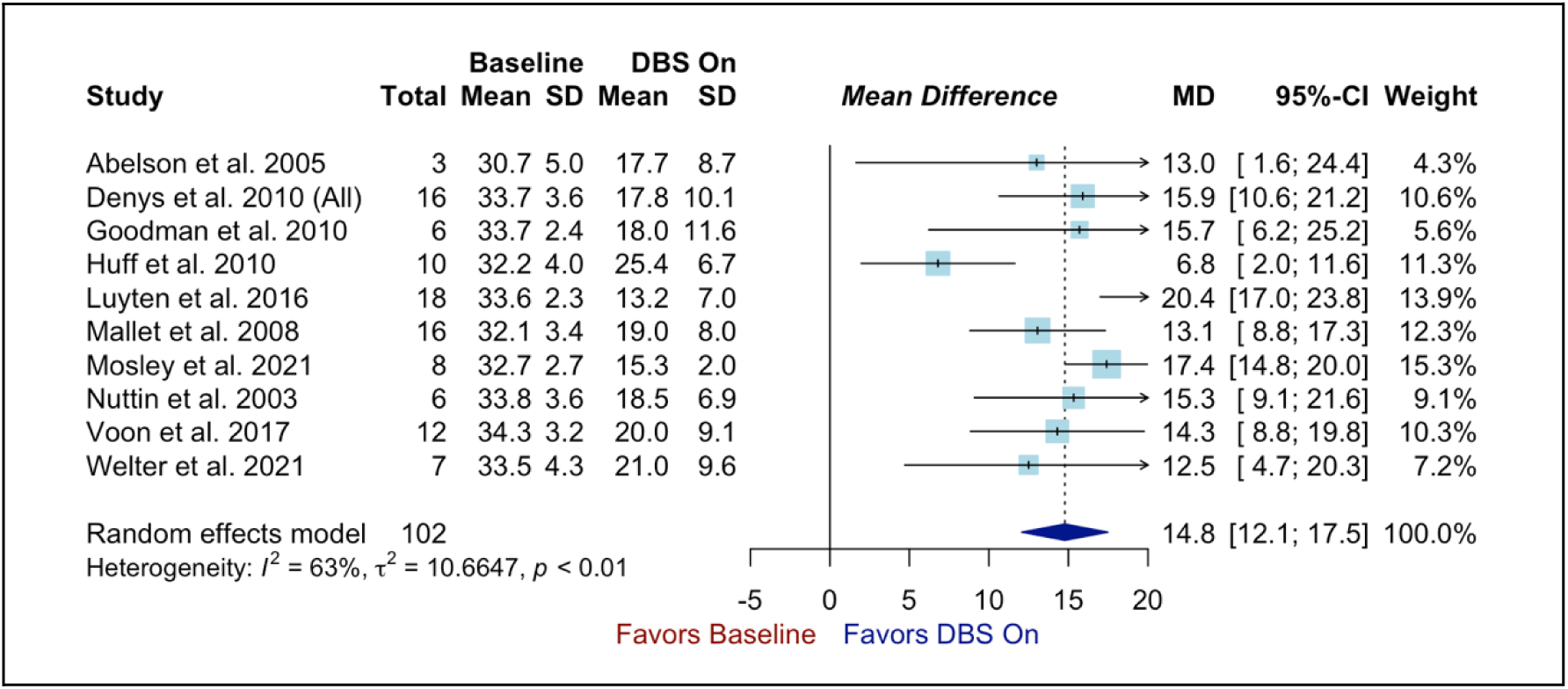
Forest plot for meta-analysis of Baseline vs. On brain stimulation. Due to the quasi-experimental nature of the evidence (before and after), the assessment started by assuming low-certainty evidence, which was raised to moderate certainty by the finding of a large intervention effect (SMD>0.8) (see SoF table).

**Table 4.**
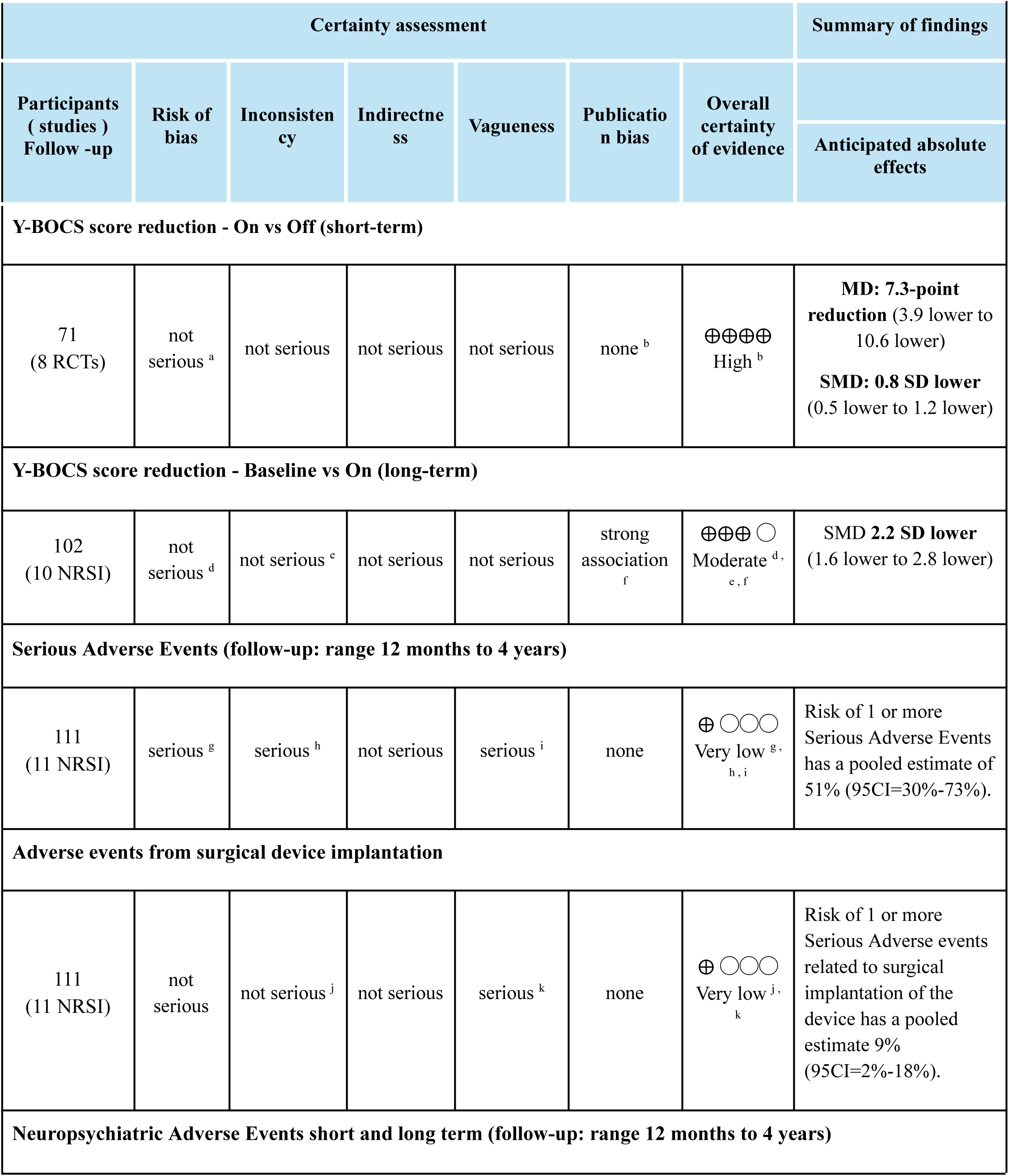
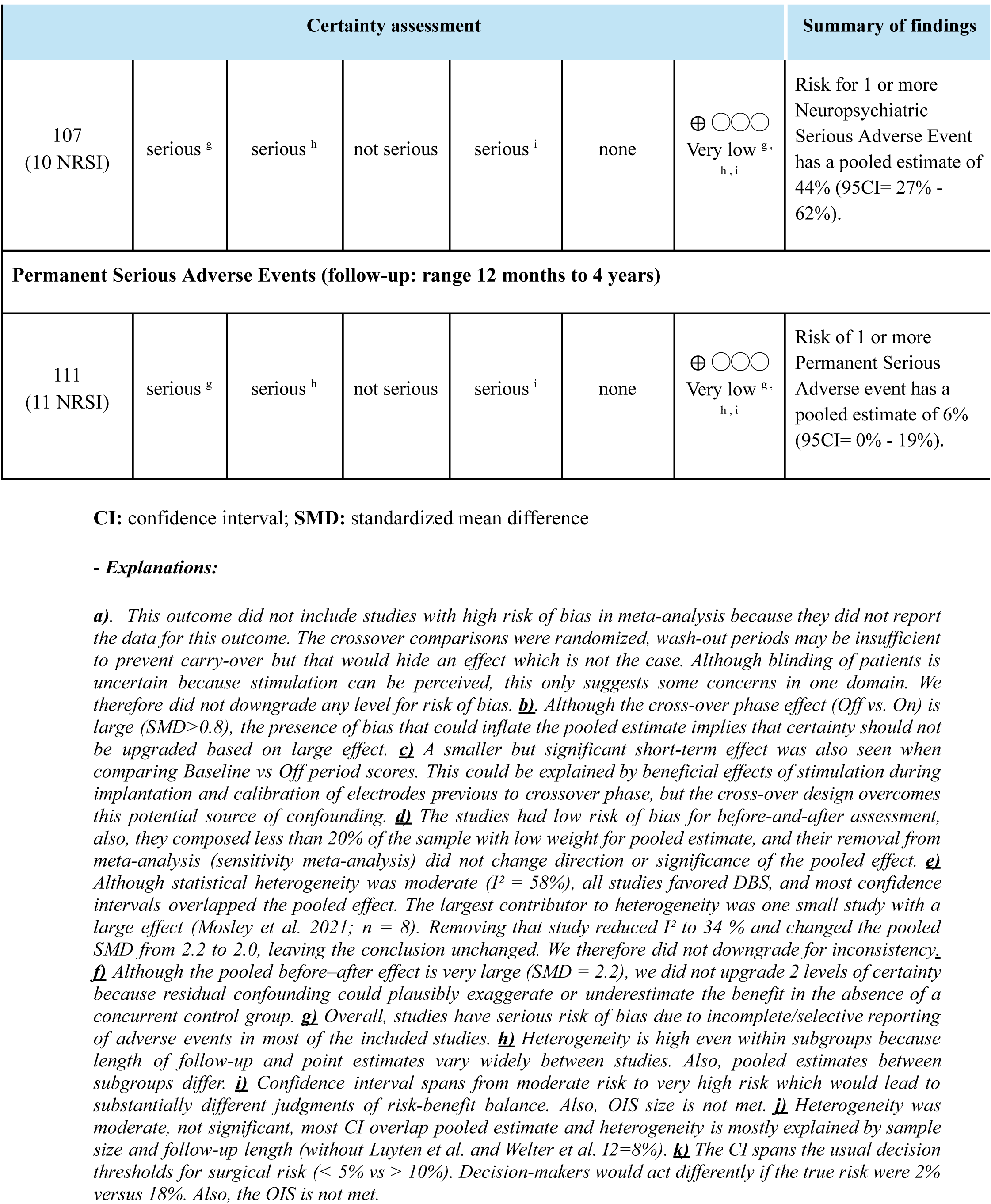
GRADE summary of findings table (SoF table)

##### Long-term Y-BOCS score reduction (Baseline vs On)

For the comparison baseline (LB) vs active stimulation (ON) long-term (>12 months) (before and after) it was possible to meta-analyze the results of 10 studies (13, 21–23, 25, 27-3 1) (n = 102 patients) regarding the effect of the intervention on the severity of OCD measured with the Y-BOCS scale . The pooled estimator for the mean difference was 14.8 (95% CI: 12.1 – 17.5) (*p* <0 . 0001) reduction in favor of DBS, with high heterogeneity (I ^2^ = 6 3 %). In the standardized mean difference (SMD) meta-analysis (see Figure S3) the effect of stimulation on the reduction in Y-BOCS score was SMD = 2.2 (95% CI: 1.6; 2.8), with high statistical heterogeneity (I 2 = 58%). However, this heterogeneity was due to two studies that showed a very large effect size of SMD = 3.8 (Luyten et al.) and SMD = 6.6 (Mosley et al.). All studies reported point estimates suggesting a benefit of the intervention, and the 95% CI of the pooled estimate only considered clinically relevant benefits. Therefore, the certainty of the evidence was not downgraded due to inconsistency. Additionally, no publication bias was detected by the Egger test (t=1.29,p=0.23, see figure S4).

### Safety of the intervention

Of the total number of studies reviewed (n=12), 10 of them analyzed the frequency of adverse events in a total of 103 patients.

Most studies reported the total number of adverse events that occurred during the study in both active and sham stimulation, except for Mallet et al (21) who collected the frequency of events in patients receiving active versus sham stimulation. Most of the studies analyzed reported the adverse events observed according to whether they were related to stimulation, implantation/surgery or the device or according to whether they were serious or not (Table 9).

#### Risk of Serious Adverse Events

Eleven studies provided evidence on the risk of serious adverse events (SAEs), involving a total of 111 patients. The pooled estimate of the risk of developing at least one SAE over the long term (12 months or more) was 51% (95% CI: 30%–73%), with high heterogeneity (I2=75%). The pooled estimates for subgroups based on follow-up time (12 months, 21 months, or 4 years) showed similar risks, so follow-up time did not explain the heterogeneity in this measure (see Figure 7). The heterogeneity in the frequency of adverse events was due to the variability in SAE definitions, active search, and reporting across studies. These factors also contributed to a serious risk of bias for this outcome. Therefore, the certainty of the evidence regarding the incidence of at least one long-term serious adverse event was very low (see Table SoF). To better understand the safety of the intervention, we assessed the risk of adverse events by type (surgical vs. neuropsychiatric) and severity, measured by permanence or irreversibility (risk of permanent AEs). The risk estimates for surgical, neuropsychiatric, and permanent AEs also have very low certainty.

**Figure 7.**
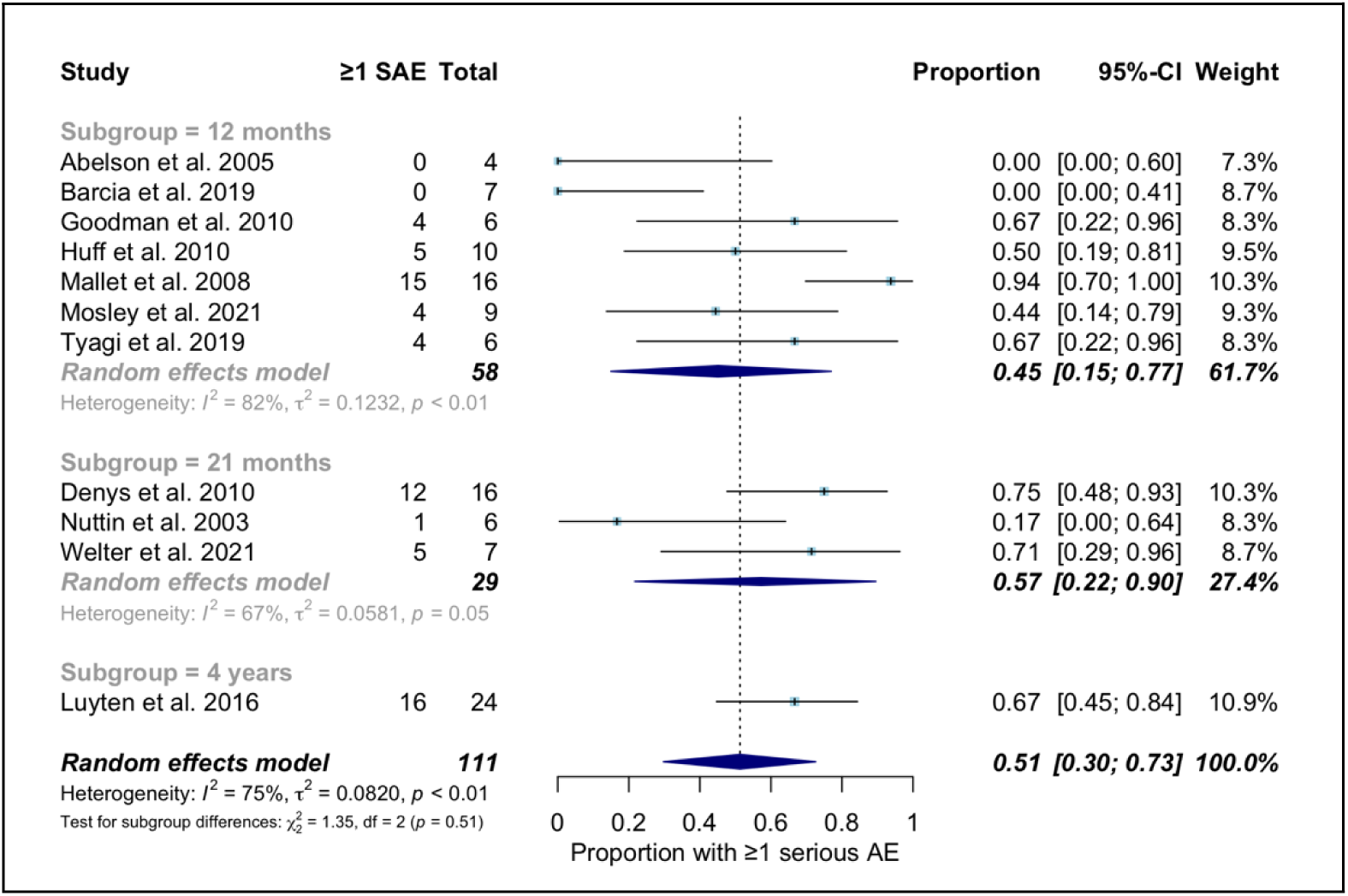
**Forest plot of meta-analysis of proportions for the risk of developing one or more SADs in the long term**

#### Risk of Serious Adverse Events from the Surgical Procedure

Eleven studies with a total of 111 patients provided evidence for this outcome . The pooled estimate of the risk of surgical SAEs was 9% (95% CI: 2%–18%) with moderate heterogeneity (I2=36%) (see Figure 8).

**Figure 8.**
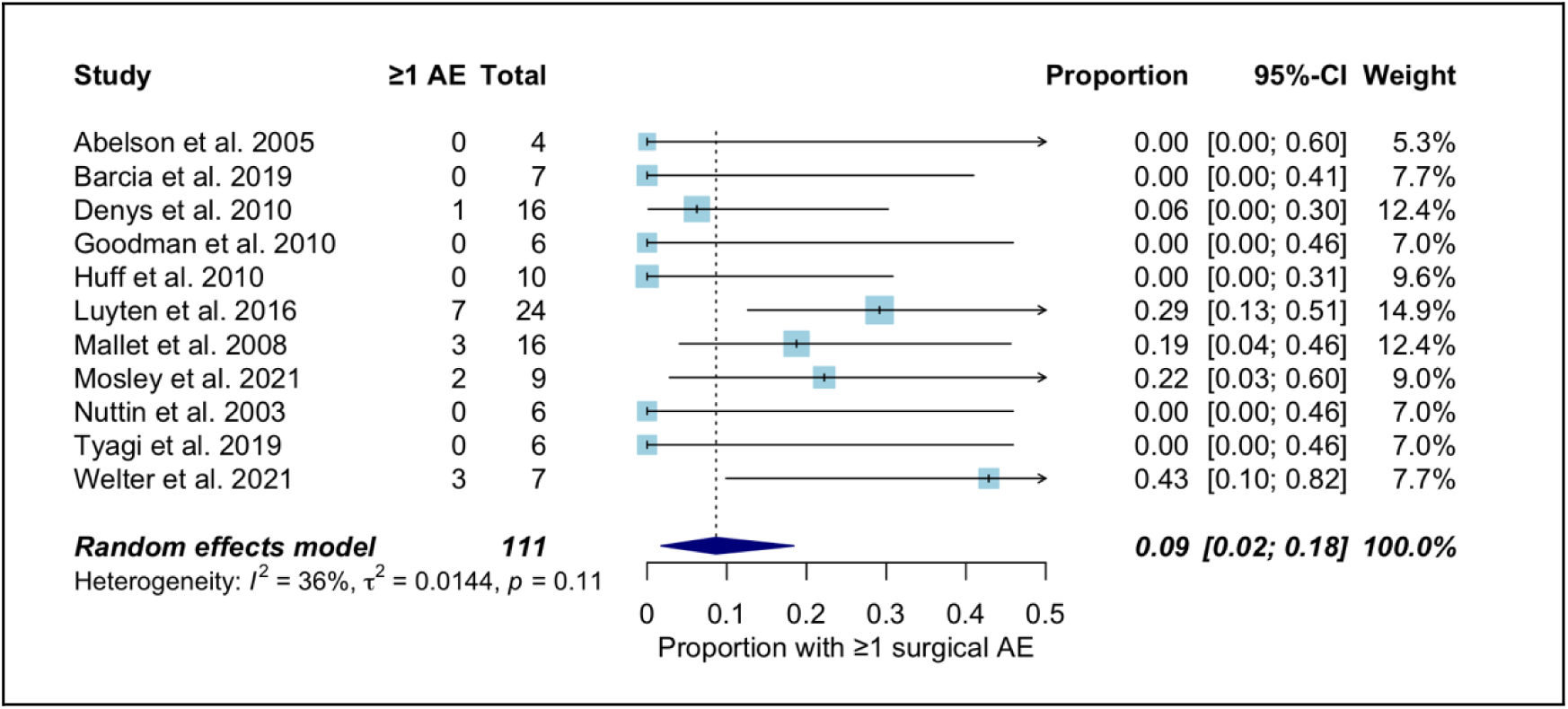
**Forest plot of meta-analysis of proportions for the risk of experiencing 1 or more surgical SAEs.**

Heterogeneity in the incidence of reported surgical SAEs was related to the sample size of the studies, as studies with small sample sizes tended to report a risk of 0%, while larger studies accounted for the majority of reported surgical SAEs .

#### Risk of Serious Adverse Events neuropsychiatric

Ten studies with a total of 107 patients provided evidence for this outcome. The pooled estimate of the risk of developing at least one neuropsychiatric SAE was 44% (95% CI: 27%–62%), with high heterogeneity (I2 = 62%), explained by the same reasons described for general SAEs (see Figure 9).

**Figure 9.**
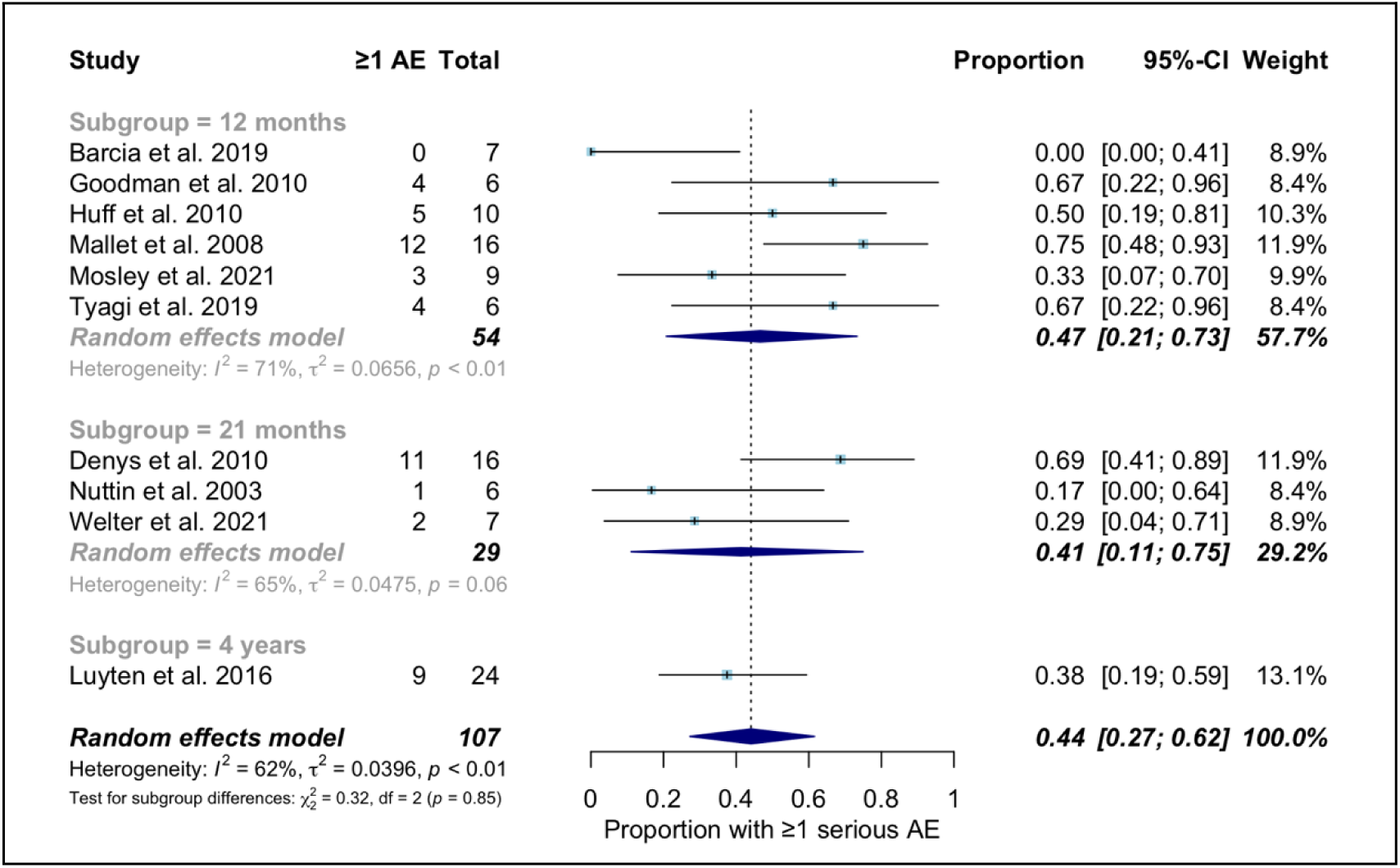
**Forest plot of meta-analysis of proportions for the risk of suffering from 1 or more neuropsychiatric SAEs**

#### Risk of Serious Adverse Events Permanent

Eleven studies with a total of 111 patients provided evidence for this outcome. The estimated pooled incidence of permanent SAEs was 6% (95% CI: 0%–19%), with high heterogeneity (I2 = 66%), explained by the same reasons described for general SAEs (see Figure 10).

**Figure 10.**
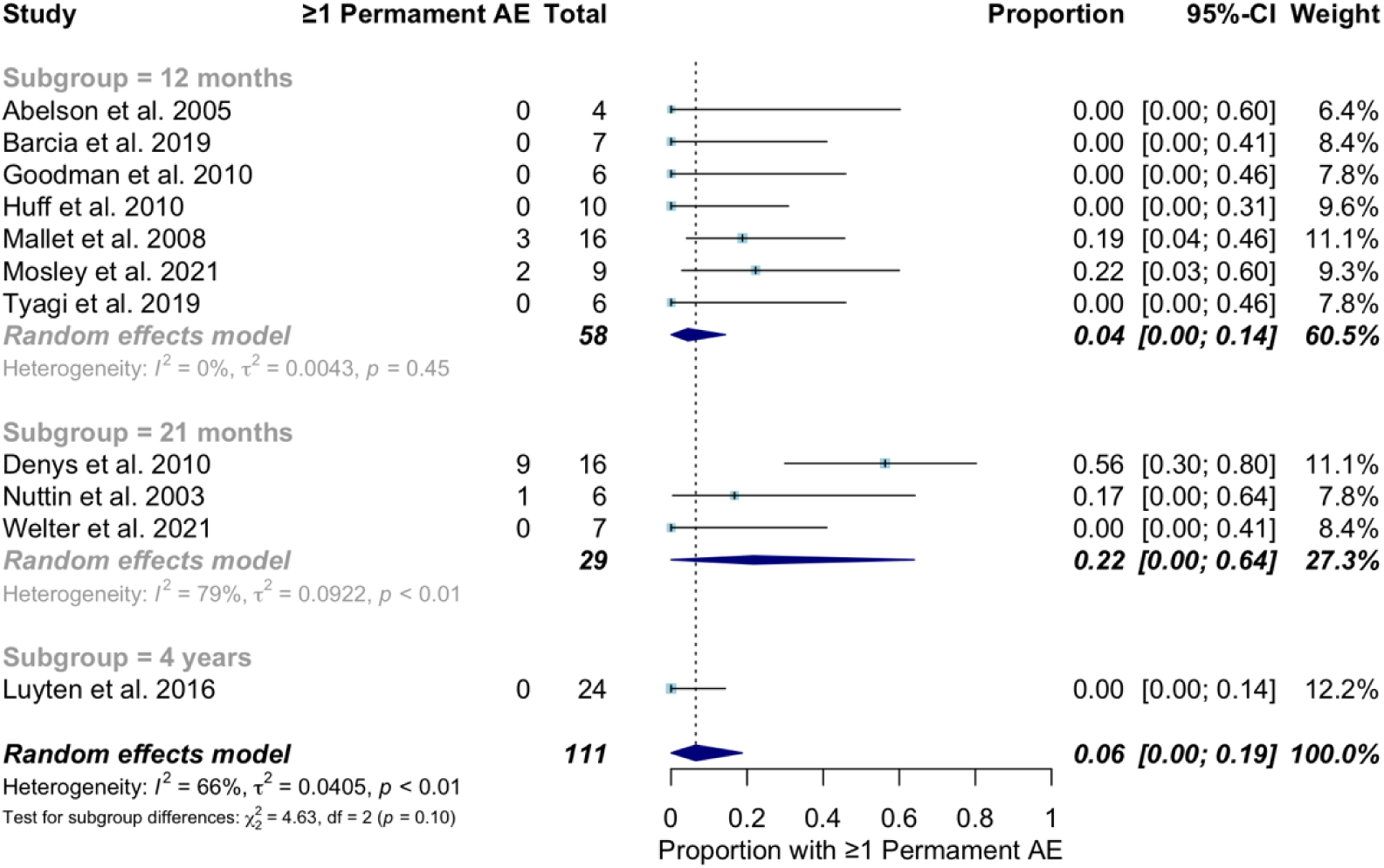
**Forest plot of meta-analysis of proportions for the risk of suffering 1 or more permanent SAEs**

Outcomes reported by individual studies – Narrative synthesis

#### CGI-S scale

Two studies reported results from this scale. Pathology severity based on clinician judgment was measured by Mallet 2008 (21) using the CGI-S scale, while Nuttin 2003 (29) presented values for the CGI-S and CGI-I subscales. In the study by Mallet et al. (reference), the CGI-S score (with lower scores indicating less disease severity) was significantly lower at the end of active stimulation than at the end of sham stimulation (p = 0.008). In the study by Nuttin (29), CGI scores remained unchanged in one patient and improved in the other three in the stimulation-on condition (See Table S5 detailing these findings) .

#### Quality of life

Quality of life was measured only in the Goodman 2010 study (13), using the SF-36 questionnaire. The authors found that only the SF-36-V item (Survey Vitality Subscale: Vigor-Activity, Fatigue-Inertia, and Vitality) showed a significant difference in the analysis of variance for repeated measures obtained during the first 12 months of DBS activation (p= 0.079). The rest of the items did not show significant differences between the ON and OFF groups.

#### Functionality

The measurement of functioning using the GAF scale (where higher scores indicate higher levels of functioning) was only used in the study by Mallet 2008 (21) where a significantly higher score was found after active stimulation than after sham stimulation (mean score at the end of active stimulation, 56 ± 14 vs 43 ± 8; p = 0.005) (Table 10). (See Table S)

#### Cognitive function

The effect of DBS on cognitive function was assessed in six studies. The domains assessed and the tasks performed varied across the included studies. Overall, the main domains assessed were memory, attention, executive function, cognitive flexibility, verbal fluency, and intellectual ability (see Table S).

Most studies did not find a relevant and/or significant effect of DBS on cognitive function (13, 21, 24, 30) . One study found that only ON DBS in amSTN (not in ECV/VS) significantly improved cognitive flexibility (26) compared to baseline and another found that active stimulation of the subthalamic nucleus increased decisional impulsivity compared to sham stimulation (22).

#### Patient perceptions and values (satisfaction, perspectives and experiences)

No included studies reported outcomes related to patient perceptions and values. Some studies collected safety outcomes related to patient-perceived discomfort and are therefore included in the safety section.

### Certainty of the evidence

The assessment of the quality of evidence for each of the outcome variables considered is presented in Table 4.

## DISCUSSION

### Summary of findings

This baseline synthesis provides an updated summary of the evidence on the use of deep brain stimulation (DBS) in patients with severe and treatment-resistant obsessive-compulsive disorder (OCD). The metanalyses revealed that DBS has a notable reduction in symptom severity in the short (high certainty) and long term (moderate certainty), as measured by the Yale-Brown Obsessive-Compulsive Scale (Y-BOCS), with pooled mean differences of 7.3 and 14.8 points depending on the time frame assessed. Nonetheless, evidence regarding the safety of the intervention was of very low certainty. Although incidence of serious and permanent adverse events was low (6%), current estimates may vary widely with future research. This uncertainty on the weighting of benefits and harms of the intervention is to be considered. To note, a high incidence of serious neuropsychiatric adverse events occurred with active stimulation or discontinuation of it during the short term follow-up periods, but were resolved with parameters adjustment or, rarely, device removal.

### Limitations

The total amount of patients included can be considered low, although it reached the OIS for the efficacy outcomes, it was insufficient for safety events, and all dichotomous outcomes for that matter. It is worth noting that the evidence related to outcomes such as quality of life and functionality was particularly scarce (derived from only one study), and none of the studies evaluated analyzed patient satisfaction or perspectives.

### Applicability of the evidence

Our results must be interpreted in consideration of the variations in the apllication of the intervention between studies, such as the methods for target selection, optimal contact identification and calibration of stimulation parameters. This unstandardized aspects of the intervention may respond to different apporaches preferred by the treating physicians, to patients’ brain anatomical and functional particularities, and to the exploration of better therapeutic targets within studies and between studies.

### Comparison with other similar publications

The main results of this review globally point in the same direction as those obtained by previous systematic reviews, both in terms of effectiveness and safety. The systematic review by Gadot et al. 2021 (2), which included all types of designs, provides results for a total sample of 352 patients with severe and resistant OCD (9 RCTs, n = 97, and 25 non-RCTs, n = 255). This review found a reduction of 14.3 points on the Y-BOCS scale, and a response rate at the last follow-up of 66% of patients classified as responders. Approximately 70% of the studies (24 of 34; n = 249) reported information on serious adverse events, which included *hardware-related complications*, infections, seizures, suicide attempts, intracranial hemorrhage, and the development of new obsessions associated with stimulation. Approximately 31% of patients (n = 78) experienced a serious adverse event. Mar-Barrutia et al (3), who also included all types of designs, analyzed the effect of short-term (mean follow-up period of 18.5 ± 8.0 months) and long-term studies (mean follow-up period was 18.5 ± 8.0 months for short-term studies and 63.7 ± 20.7 months). In this review, a mean reduction in the Y-BOCS score of 47.4% ± 21% was found in short-term studies and 47.2% ± 9.9% in long-term studies. The response to stimulation was stable over time, with a greater number of patients meeting response criteria in long-term studies (70.7%) than in short-term studies (60.6%). No differences in safety were found.

In short-term vs. long-term studies, most adverse events were mild and transient, and improved after adjusting stimulation parameters. Martinho et al (20) also found that DBS decreased the severity of symptoms, but with a relevant presence of adverse events and dropouts. These results are consistent with our findings in terms of the low possibility of a serious adverse events such as (32, 33) .

### Clinical implications

Overall, the results indicate that this technique should only be performed in carefully selected cases, after failure of all recommended therapeutic options for the management of OCD and after individualized assessment of the benefits and risks of the technique through a process of shared decision-making and appropriate informed consent. Benefit with proposed protocols is likely to be perceive and is likely to be large. Although estimates of SAE incidence are uncertain, they can be drawn from the risks described for neurosurgical procedures of electrode implantation for long-term use. Balance of benefits and harms is to be discussed with patients according to their individual clinical characteristics and preferences.

### Implications for research

Further studies on the effectiveness and safety of DBS in patients with severe and treatment-resistant OCD are needed. Specifically, comparative effectiveness studies of different interventions targeting severe, treatment-resistant OCD, such as transcranial magnetic stimulation or ablative surgery, would aid in decision-making regarding the most appropriate intervention for each patient. Furthermore, it is important to generate scientific evidence to delve deeper into the significant uncertainties surrounding therapeutic targets, optimal stimulation protocols, and the personalization or individualization of the intervention, and minimization of adverse events.

Studies should employ standardized instruments for assessing severity (Y-BOCS, CGI), measuring impact on quality of life (e.g., SF-36) and functionality (GAF), and assessing outcomes related to the patient perspective (acceptability, satisfaction, values, and preferences), in addition to comprehensively reporting on potential adverse events associated with the intervention.

### Living evidence approach

This synthesis is part of a larger project designed to implement a living evidence framework. This project aims to produce multiple, parallel, relevant living systematic reviews to inform decision-making, following the highest quality standards for producing evidence syntheses. This framework is well-suited to integrating emerging scientific evidence on the role of DBS in severe and refractory OCD. We have identified several ongoing studies addressing this issue, which will provide valuable evidence to inform decision-makers in the near future. In The Living Evidence to Inform Health Decisions website (https://livingevidenceframework.com/en/lesr/) a living version of this synthesis will be available and we will resubmit the review for publication whenever there are substantial updates in certainty of evidence and the conclusions change.

## Data Availability

All data produced in the present study are available upon reasonable request to the authors

## Data Availability

All data produced in the present study are available upon reasonable request to the authors

## Acknowledgments

To the Living Evidence to Inform Health Decisions Program research group (Maria X. Rojas, Gerard Urrutia; Ariadna Auladell; Josefina Bendersky; Laura Trujillo; Luz Angela Torres) for their contribution to the design, planning and review of this SR.

To the the methodological team of the Epistemonikos Foundation to the development of this review.

## Roles and contributions

The idea was conceived by Yolanda Triñanes Pego, Patricia Gómez Salgado, Janet Puñal Riobóo, and María del Carmen Maceira Rozas, who also developed the protocol. María Ximena Rojas provided advice on defining the living evidence synthesis approach. The original draft of the manuscript was prepared by Yolanda Triñanes and Patricia Gómez Salgado. Andrés Gempelers contributed methodological advice and support, conducted risk of bias assessments, performed meta-analyses, assessed the certainty of evidence, and made important intellectual contributions to the final manuscript. María José Faraldo Vallés contributed to the conception of the idea, the development of the protocol, and the internal review of the document. All authors contributed to the drafting and final approval of the manuscript.

## Financing

The evaluation of the effectiveness and safety of DBS in resistant OCD has been commissioned to the Galician Agency for Knowledge in Health by the Advisory Commission for the Incorporation of Techniques, Technologies or Procedures into the Service Galician Health .

This ongoing work is being developed as part of the Living Evidence to Inform Health Decisions (LE-IHD) program and funded by the Instituto de Salud Carlos III (ISCIII) (Grant PI21/01564) througth the project “Strengthening decision-making capacity in the Spanish Health System through living evidence: An innovative framework,” and co-funded by the European Union through FEDER and the Horizon’s 2020 research and innovation programme (Grant Agreement MSCA-IF-EF-ST #894990), awarded to María Ximena Rojas for the development of the Living Evidence to Inform Health Decisions project.

The funding institution played no role in the study design, the drafting, review, and approval of the protocol, or the decision to submit it for publication. The LE-IHD program provided training, support, and tools at no cost to support the development of this protocol.

## Ethical aspects

Since the researchers will not have access to information that could lead to the identification of participants, it was not necessary to obtain approval from an ethics committee.

## Bibliographic references

1. American Psychiatric Association. Diagnostic and Statistical Manual of Mental Disorders (DSM-5®), 5th Ed. Arlington, VA, American Psychiatric Association. 2014. [accessed 13 Nov 2023]. Available at: https://www.federaciocatalanatdah.org/wp-content/uploads/2018/12/dsm5-manualdiagnsticoyestadisticodelostrastornosmentales-161006005112.pdf.

2. Gadot R, Najera R, Hirani S, Anand A, Storch E, Goodman WK, et al. Efficacy of deep brain stimulation for treatment-resistant obsessive-compulsive disorder: systematic review and meta-analysis. Journal of Neurology, Neurosurgery and Psychiatry. 2022. PubMed PMID: 36127157.

3. Mar-Barrutia L, Real E, Segalas C, Bertolin S, Menchon JM, Alonso P. Deep brain stimulation for obsessive-compulsive disorder: A systematic review of worldwide experience after 20 years. World J Psychiatry. 2021;11(9):659–80. PubMed PMID: 34631467.

4. Fineberg NA, Hollander E, Pallanti S, Walitza S, Grunblatt E, Dell’Osso BM, et al. Clinical advances in obsessive-compulsive disorder: a position statement by the International College of Obsessive-Compulsive Spectrum Disorders. International Clinical Psychopharmacology. 2020;35(4):173–93. PubMed PMID: 32433254.

5. Katzman MA, Bleau P, Blier P, Chokka P, Kjernisted K, Van Ameringen M, et al. Canadian clinical practice guidelines for the management of anxiety, posttraumatic stress and obsessive-compulsive disorders. BMC Psychiatry. 2014;14 Suppl 1(Suppl 1):S1. PubMed PMID: 25081580.

6. Menchon JM, Bobes J, Alamo C, Alonso P, Garcia-Portilla MP, Ibanez A, et al. Pharmacological treatment of obsessive compulsive disorder in adults: A clinical practice guideline based on the ADAPTE methodology. Rev Psiquiatr Salud Ment (Engl Ed). 2019;12(2):77–91. PubMed PMID: 30850318.

7. Nezgovorova V, Reid J, Fineberg NA, Hollander E. Optimizing first line treatments for adults with OCD. Comprehensive Psychiatry. 2022;115:152305. PubMed PMID: 35325671.

8. Van Ameringen M, Simpson W, Patterson B, Dell’Osso B, Fineberg N, Hollander E, et al. Pharmacological treatment strategies in obsessive compulsive disorder: A cross-sectional view in nine international OCD centers. J Psychopharmacol. 2014;28(6):596–602. PubMed PMID: 24429223.

9. Roh D, Jang KW, Kim CH. Clinical Advances in Treatment Strategies for Obsessive-compulsive Disorder in Adults. Clin Psychopharmacol Neurosci. 2023;21(4):676–85. PubMed PMID: 37859440.

10. CORDIS EU research results. Living Evidence to inform health decisions: Development of capacity for the production and use of reliable, friendly and updated health evidence synthesis. Available at: https://cordis.europa.eu/project/id/894990. Cited: December 2023.

11. Rethlefsen ML, Kirtley S, Waffenschmidt S, Ayala AP, Moher D, Page MJ, et al. PRISMA-S: an extension to the PRISMA statement for reporting literature searches in systematic reviews. Journal of the Medical Library Association. 2021;109(2):174–200. PubMed PMID: 34285662.

12. Moher D, Shamseer L, Clarke M, Ghersi D, Liberati A, Petticrew M, et al. Preferred reporting items for systematic review and meta-analysis protocols (PRISMA-P) 2015 statement. Syst Rev 2015 Jan 1;4(1):1. PubMed PMID: 25554246.

13. Goodman WK, Foote KD, Greenberg BD, Ricciuti N, Bauer R, Ward H, et al. Deep brain stimulation for intractable obsessive compulsive disorder: pilot study using a blinded, staggered-onset design. Biological Psychiatry. 2010;67(6):535–42. PubMed PMID: 20116047.

14. Bendersky J, Auladell-Rispau A, Urrútia G, Rojas-Reyes MX. Methods for developing and reporting living evidence synthesis. Journal of Clinical Epidemiology. 2022 Dec;152:89–100. PubMed PMID: 36220626.

15. Epistemonikos Foundation. L·OVE platform. https://iloveevidence.com/.

16. Shea BJ, Reeves BC, Wells G, Thuku M, Hamel C, Moran J, et al. AMSTAR 2: a critical appraisal tool for systematic reviews that include randomized or non-randomised studies of healthcare interventions, or both. BMJ. 2017;358:j4008. PubMed PMID: 28935701.

17. Cochrane Scientific Committee. Risk of Bias 2 (RoB 2) tool [Internet]. London: Cochrane; 2020 [cited March 10, 2023]. Available at: https://methods.cochrane.org/risk-bias-2.

18. Guyatt GH, Oxman AD, Vist GE, Kunz R, Falck-Ytter Y, Alonso-Coello P, et al. GRADE: an emerging consensus on rating quality of evidence and strength of recommendations. BMJ. 2008 Apr 26;336(7650):924–6. PubMed PMID: 18436948.

19. The GRADE Working Group. Grading of Recommendations Assessment, Development and Evaluation (short GRADE) [Internet]. 2021 [cited Feb 27, 2021]. Available at: https://www.gradeworkinggroup.org/.

20. Martinho FP, Duarte GS, Couto FSD. Efficacy, Effect on Mood Symptoms, and Safety of Deep Brain Stimulation in Refractory Obsessive-Compulsive Disorder: A Systematic Review and Meta-Analysis. Journal of Clinical Psychiatry. 2020;81(3). PubMed PMID: 32459406.

21. Mallet L, Polosan M, Jaafari N, Baup N, Welter ML, Fontaine D, et al. Subthalamic nucleus stimulation in severe obsessive-compulsive disorder. New England Journal of Medicine. 2008;359(20):2121–34. PubMed PMID: 19005196.

22. Voon V, Droux F, Morris L, Chabardes S, Bougerol T, David O, et al. Decisional impulsivity and the associative-limbic subthalamic nucleus in obsessive-compulsive disorder: stimulation and connectivity. Brain. 2017;140(2):442–56. PubMed PMID: 28040671.

23. Welter M, Flores Alves dos Santos J, Clair A, Lau B, Mamadou Diallo H, Fernandez-Vidal S, et al. Deep Brain Stimulation of the Subthalamic, Accumbens, or Caudate Nuclei for Patients With Severe Obsessive-compulsive Disorder: A Randomized Crossover Controlled Study. Biological Psychiatry. 2021;90(10):pp.e45–e7.

24. Barcia JA, Avecillas-Chasin JM, Nombela C, Arza R, Garcia-Albea J, Pineda-Pardo JA, et al. Personalized striatal targets for deep brain stimulation in obsessive-compulsive disorder. Brain Stimulation. 2019;12(3):724–34. PubMed PMID: 30670359.

25. Huff W, Lenartz D, Schormann M, Lee SH, Kuhn J, Koulousakis A, et al. Unilateral deep brain stimulation of the nucleus accumbens in patients with treatment-resistant obsessive-compulsive disorder: Outcomes after one year. Clinical Neurology and Neurosurgery. 2010 Feb;112(2):137–43. PubMed PMID: 20006424.

26. Tyagi H, Apergis-Schoute AM, Akram H, Foltynie T, Limousin P, Drummond LM, et al. A Randomized Trial Directly Comparing Ventral Capsule and Anteromedial Subthalamic Nucleus Stimulation in Obsessive-Compulsive Disorder: Clinical and Imaging Evidence for Dissociable Effects. Biological Psychiatry. 2019;85(9):726–34. PubMed PMID: 30853111.

27. Denys D, Mantione M, Figee M, van den Munckhof P, Koerselman F, Westenberg H, et al. Deep brain stimulation of the nucleus accumbens for treatment-refractory obsessive-compulsive disorder. Archives of General Psychiatry. 2010;67(10):1061–8. PubMed PMID: 20921122.

28. Luyten L, Hendrickx S, Raymaekers S, Gabriels L, Nuttin B. Electrical stimulation in the bed nucleus of the stria terminalis alleviates severe obsessive-compulsive disorder. Molecular Psychiatry. 2016;21(9):1272–80. PubMed PMID: 26303665.

29. Nuttin BJ, Gabriels LA, Cosyns PR, Meyerson BA, Andreewitch S, Sunaert SG, et al. Long-term electrical capsular stimulation in patients with obsessive-compulsive disorder. Neurosurgery. 2003;52(6):1263–72; discussion 72-4. PubMed PMID: 12762871.

30. Abelson J, Curtis G, Sagher O, Albucher R, Harrigan M, Taylor S, et al. Deep Brain Stimulation for Refractory Obsessive–Compulsive Disorder. BIOL PSYCHIATRY 2005;57:510–6.

31. Mosley PE, Windels F, Morris J, Coyne T, Marsh R, Giorni A, et al. A randomized, double-blind, sham-controlled trial of deep brain stimulation of the bed nucleus of the stria terminalis for treatment-resistant obsessive-compulsive disorder. Transl Psychiatry. 2021;11(1):190. PubMed PMID: 33782383.

32. Alonso P, Cuadras D, Gabriëls L, Denys D, Goodman W, Greenberg BD, et al. Deep Brain Stimulation for Obsessive-Compulsive Disorder: A Meta-Analysis of Treatment Outcome and Predictors of Response. PloS One. 2015;10(7):e0133591. PubMed PMID: 26208305.

33. Kisely S, Hall K, Siskind D, Frater J, Olson S, Crompton D. Deep brain stimulation for obsessive-compulsive disorder: a systematic review and meta-analysis. Psychological Medicine. 2014 Dec;44(16):3533–42. PubMed PMID: 25066053.

34. Triñanes Y, Gómez P, Puñal J, Maceira MC, The Living Evidence to Inform Health Decision program (LE-IHD) research group. Effectiveness and safety of deep brain stimulation in severe and treatment-resistant obsessive-compulsive disorder (Protocol). 2023. [accessed. Available at: 10.17605/OSF.IO/RBT98 .

